# Impact of Androgenic Anabolic Steroids on Cardiovascular Health in Men and Women - PART OF THE FITNESS DOPING IN DENMARK (FIDO-DK) STUDY

**DOI:** 10.1101/2024.11.18.24317516

**Authors:** Laust Frisenberg Buhl, Louise Lehmann Christensen, Rikke Hjortebjerg, Selma Hasific, Clara Hjerrild, Stefan Harders, Mads Lillevang-Johansen, Dorte Glintborg, Marianne S. Andersen, Mario Thevis, Caroline Kistorp, Jon Jarløv Rasmussen, Jes S. Lindholt, Axel Diederichsen, Jan Frystyk

## Abstract

**Background:** Illicit use of anabolic androgenic steroids (AAS) is common among recreational athletes, yet comprehensive studies on adverse cardiovascular outcomes, especially in female AAS users, are lacking.

**Methods:** A cross-sectional study of recreational athletes of women and men was conducted, involving active and previous AAS users and non-users aged ≥18 years. Previous use was defined as discontinuation of AAS at least three months prior to study. Primary outcomes included atherosclerosis (carotid, femoral, and coronary artery plaques) and cardiac function, assessed using vascular ultrasound, coronary computed tomography angiography and echocardiography.

**Results:** Median age was 36 years for active users (n=80, 19 women), 35 years for previous users (n=26, 8 women), and 40 years for non-users (n=58, 16 women) (p=NS). Median AAS usage period was 2.2 years for both active and previous users; the latter group had discontinued intake 2.5 years before study (range: 3 months to 29 years).

There was no group differences when comparing the number of femoral/carotid artery plaques, the coronary artery calcium (CAC) score or the number of non-calcified plaques. However, confounder-adjusted logistic regression showed associations between cumulative AAS use and a positive CAC score (OR: 1.23, 95% CI: 1.09–1.39, p=0.001) and the presence of non-calcified plaque (OR: 1.17, 95% CI: 1.05–1.30, p=0.004), respectively, when comparing previous and ongoing users vs. non-users. These associations were also present in men, but not women. Moreover, >5 years of AAS use increased the fraction of athletes with increased severity of calcifications (p=0.043). Echocardiography showed that active AAS using males and females had impaired left ventricular global longitudinal strain (LVGLS) and right ventricular global longitudinal strain (RVGLS) compared to sex-matched non-users (p<0.001). Multivariable analysis showed that cumulative AAS use correlated with worsening of LVGLS (p=0.002) and RVGLS (p=0.001). Finally, after 5 years of cumulative AAS use, nearly all athletes had ventricular mass above and left ventricular ejection fraction below the median of normal range.

**Conclusion:** In men, the cumulative lifetime AAS exposure was an independent predictor of coronary atherosclerosis. However, both male and female AAS users share risks of myocardial dysfunction, underscoring significant cardiovascular risks across genders.

**CLINICAL PERSPECTIVE:** Key observations from the study:

- In recreational athletes, the accumulated lifetime AAS exposure associates with a higher prevalence of non-calcified plaques and coronary artery calcification in male recreational athletes.
- Our study suggests that more than 5 years of AAS use constitutes a threshold beyond which the development of coronary calcifications significantly increases compared to non-users.
- In addition to compromised left ventricular systolic and diastolic function, AAS users exhibited significantly reduced right ventricular function, indicating a biventricular cardiac impact of AAS.
- Male and female AAS users showed similar patterns of cardiac deterioration.

These findings highlight the significant cardiovascular risks associated with AAS use in both male and female recreational athletes, underscoring the importance of targeted research, educational programs, information campaigns, and intervention strategies for this population, regardless of gender.

## INTRODUCTION

Originally, the illicit use of androgenic anabolic steroids (AAS) was predominantly linked to male elite athletes and bodybuilders. Today, however, recreational athletes constitute the largest group of illicit AAS users and their use of AAS is widespread, with an estimated lifetime prevalence of AAS use of 3.3% among men and 1.6% among women (**1–4**). This prevalence is concerning because long-term AAS intake has been linked to numerous detrimental health outcomes, particularly cardiovascular complications (**5**) and premature death (**6**). Notably, most of the knowledge regarding the long-term cardiovascular effects of illicit AAS use derives from studies on males, despite the fact that AAS is also used illicitly by female athletes (**3**).

The first associations between AAS use and coronary artery disease (CAD) emerged from case reports and small observational studies involving selected groups of male athletes having an illicit use of AAS. These studies reported severe premature CAD (**7–9**), myocardial infarction (**10, 11**) and sudden cardiac death (**12–14**) in younger male AAS-using bodybuilders and weightlifters (below 40 years of age). Over the last decade, a few larger studies (**9, 15–17**) have substantiated these findings, reporting a positive association between coronary artery plaque volume and the duration of AAS use (**15**). Nevertheless, there is no available data on peripheral artery disease (PAD) in this population.

Beyond CAD, substantial evidence based on post-mortem examinations (**18–20**), echocardiographic assessments (**15, 16, 21–27**), and cardiac magnetic resonance imaging (MRI) (**28, 29**) has highlighted a potential AAS-related cardiomyopathy characterized by left ventricular (LV) remodeling or hypertrophy, leading to a decline in both LV systolic and diastolic function (**15, 16, 22–29**), along with right ventricular (RV) systolic function (**30**).

To date, studies on the cardiovascular health of recreational athletes using AAS illicitly have primarily focused on myocardial deterioration in selected groups of male athletes. Consequently, there is a gap in our knowledge concerning the cardiovascular status of the broader population of recreational athletes using AAS, including females.

With these considerations in mind, it was decided to describe the cardiovascular status in a broad and unselected population of Danish AAS-using male and female recreational athletes, and to compare findings to an age- and sex-matched group of non-AAS-using recreational athletes. The primary objectives were to assess peripheral artery plaque formation through vascular ultrasound of the carotid and femoral arteries, and to evaluate coronary artery calcifications (CAC) and coronary non-calcified plaques (NCP) using coronary computed tomography angiography (CCTA). Finally, echocardiography, lifestyle, medical history, socioeconomic status, and biochemistry was included.

## METHODS

### Recruitment of participants

The Fitness Doping in Denmark (FIDO-DK) study (ClinicalTrials.gov: NCT05178537) is a nationwide cross-sectional cohort study. Here were report data from Odense University Hospital, Denmark. Our aim was to examine a broad and unselected population of recreational athletes, including both men and women aged 18 and older who had been using AAS illicitly for at least three months, without imposing any additional inclusion criteria. A detailed study protocol has been published recently (**31**).

Participants were recruited via targeted announcements, social media posts, flyers, advertisements, and posters in training centers. Interested recreational athletes received detailed information by email and telephone. Participants from various parts of Denmark were enrolled after providing informed written and oral consent. After enrolling AAS users, this group was matched with a control group of healthy non-users of similar sex and age demographics, who engaged in strength training at least twice a week. Non-users were recruited in the same manner as AAS users. According to the protocol, it was planned to enroll 120 male and female athletes with active or previous AAS use and 60 non-users and we managed to include 107 AAS users and 58 non-users (**Table 1**).

**Table 1.** Characteristics of the participants in the entire cohort stratified by AAS use. Values for continuous variables are shown as median (interquartile range). Categorical variables are indicated as number (n) and percentage (%) of persons. *p<0.05 when compared to non-users, ^†^p<0.05 when compared to previous AAS users. #p<0.05 when comparing men and women within the same group. This comparison was only done for the cumulative use of AAS. AAS, androgenic anabolic steroids; BMI, body mass index.

| Variable | Active AAS users<br>(n=80) | Previous AAS users<br>(n=26) | Non-users<br>(n=58) | P-value |
| --- | --- | --- | --- | --- |
| Male sex, n (%) | 61 (76) | 18 (69) | 42 (72) | 0.62 |
| Age, years | 35 (30;43) | 36 (28;51) | 40 (31;46) | 0.14 |
| - Men | 33 (28;39)<sup>*</sup> | 35 (28;49) | 43 (31;50) | **0.019** |
| - Women | 44 (33;47) | 39 (32;53) | 36 (32;43) | 0.21 |
| Height, m | 178 (171;184)<sup>*</sup> | 175 (169;181)<sup>*</sup> | 182 (175;186) | **0.018** |
| - Men | 181 (175;186)<sup>*</sup> | 179 (172;183)<sup>*</sup> | 183 (182;188) | **0.006** |
| - Women | 165 (161;171)<sup>*</sup> | 170 (161;175) | 173 (169;175) | 0.036 |
| Body surface area, m<sup>2</sup> | 2.10 (1.92;2.29) | 1.99 (1.87;2.15) | 2.07 (1.93;2.14) | 0.14 |
| - Men | 2.17 (2.06;2.33) | 2.06 (1.93;1.79) | 2.11 (2.05;2.24) | 0.098 |
| - Women | 1.81 (1.73;1.92) | 1.79 (1.72;1.92) | 1.84 (1.74;1.93) | 0.81 |
| BMI, kg/m<sup>2</sup> | 29.0 (26.4;31.4)<sup>*,†</sup> | 26.5 (25.0;28.6) | 25.2 (23.4;27.9) | **<0.001** |
| - Men | 29.4 (27.7;32.2)<sup>*,†</sup> | 27.4 (25.6;29.9) | 26.5 (24.0;28.3) | **<0.001** |
| - Women | 25.9 (24.9;29.1)<sup>*</sup> | 24.6 (22.9;26.3) | 24.3 (22.9;25.5) | **0.036** |
| Fat percentage, % | 14.8 (11.1;17.2)<sup>*,†</sup> | 18.2 (14.2;22.2) | 21.4 (17.7;23.6) | **<0.001** |
| - Men | 13.1 (10.4;16.5)<sup>*,†</sup> | 17.8 (12.3;21.2) | 20.0 (16.3;22.5) | **<0.001** |
| - Women | 16.5 (15.9;21.6)<sup>*</sup> | 20.2 (16.7;26.9) | 25.5 (22.1;27.9) | **0.006** |
| Muscle percentage, % | 41.3 (38.4;43.1)<sup>*,†</sup> | 38.4 (34.1;41.1) | 37.2 (33.9;40.0) | **<0.001** |
| - Men | 41.7 (39.8;43.6)<sup>*</sup> | 40.0 (37.8;42.7) | 38.7 (36.3;40.4) | **<0.001** |
| - Women | 36.6 (31.9;41.5)<sup>*,†</sup> | 33.8 (30.4;34.5) | 32.2 (29.8;34.9) | **0.007** |
| Systolic blood pressure, mmHg | 138 (129;147)<sup>*,†</sup> | 123 (110;131)<sup>*</sup> | 132 (125;139) | **<0.001** |
| - Men | 138 (130;148)<sup>*,†</sup> | 129 (122;134) | 135 (126;141) | **0.002** |
| - Women | 135 (116;143)<sup>†</sup> | 107 (104;113)<sup>*</sup> | 128 (121; 126) | **<0.001** |
| Diastolic blood pressure, mmHg | 77 (71;84)<sup>†</sup> | 72 (68;75)<sup>*</sup> | 78 (68;83) | **0.016** |
| - Men | 77 (70;84) | 75 (71;78) | 78 (68;82) | 0.34 |
| - Women | 77 (71;83)<sup>†</sup> | 68 (68;69)<sup>*</sup> | 78 (71;86) | **0.005** |
| Time spent on hard exercise and strength training, hours | 11 (8.5;17)<sup>*</sup> | 10 (5.0;14)<sup>*</sup> | 5.0 (4.0;10) | **<0.001** |
| - Men | 11 (7;17)<sup>*</sup> | 10 (5.0;13.5)<sup>*</sup> | 5.0 (2.0;10) | **<0.001** |
| - Women | 10 (10;17)<sup>*</sup> | 12 (6.0;17)<sup>*</sup> | 4.0 (4.0;7.0) | **<0.001** |
| Hand grip strength (UNIT) | 58 (48;65)<sup>*,†</sup> | 51 (39;59) | 53 (43;62) | **0.006** |
| - Men | 61 (55;70)<sup>*,†</sup> | 57 (50;61) | 58 (51;64) | **0.006** |
| - Women | 40 (37;44) | 35 (32;38) | 40 (32;42) | 0.17 |
| Maximal bench press weight, kg | 120 (100;153)<sup>*,†</sup> | 100 (60;120) | 90 (78;110) | **<0.001** |
| - Men | 140 (118;165)<sup>*,†</sup> | 110 (95;125) | 100 (90;120) | **<0.001** |
| - Women | 84 (50;95)<sup>†</sup> | 35 (30;60) | 49 (30;60) | **0.049** |
| Family history of coronary disease, n (%) | 15 (19.5) | 3 (12.0) | 5 (8.8) | 0.20 |
| - Men | 8 (13.3) | 3 (17.7) | 2 (4.9) | 0.22 |
| - Women | 7 (41.2) | 0 (0.0) | 3 (18.8) | 0.073 |
| Cumulative lifetime total duration of AAS use, years | 2.2 (1.2;7.2)<sup>*</sup> | 2.2 (1.0;5.5)<sup>*</sup> | 0 (0;0) | **<0.001** |
| - Men | 3.8 (1.3;9.8)<sup>*</sup> | 2.4 (1.2;5.5)<sup>*</sup> | 0 (0;0) | **<0.001** |
| - Women | 2.0 (0.5;2.8)<sup>*,#</sup> | 1.4 (0.4;3.9)<sup>*,#</sup> | 0 (0;0) | **<0.001** |
| Smoking status |  |  |  |  |
| Never | 32 (41.3)<sup>*</sup> | 12 (46.2) | 39 (67.2) | **0.017** |
| Previous | 21 (26.9) | 9 (34.6) | 12 (20.7) |  |
| Current | 25 (32.1) | 5 (19.2) | 7 (12.1) |  |
| - Men |  |  |  | **0.040** |
| Never | 25 (41.7)<sup>*</sup> | 8 (44.4) | 29 (69.1) |  |
| Previous | 16 (26.7) | 5 (27.8) | 9 (21.4) |  |
| Current | 19 (31.7) | 5 (27.8) | 4 (9.5) |  |
| - Women |  |  |  | 0.25 |
| Never | 7 (38.9) | 4 (50.0) | 10 (62.5) |  |
| Previous | 5 (27.8) | 4 (50.0) | 3 (18.8) |  |
| Current | 6 (33.3) | 0 (0.0) | 3 (18.8) |  |
| Weekly alcohol consumption, n (%) |  |  |  |  |
| 0 units | 46 (59.0)<sup>*</sup> | 14 (52.9)<sup>*</sup> | 8 (13.8) | **<0.001** |
| 1-7 units | 30 (38.5) | 11 (42.3) | 40 (69.0) |  |
| 8-14 units | 2 (2.6) | 1 (3.9) | 10 (17.2) |  |
| ≥14 units | 0 (0.0) | 0 (0.0) | 0 (0.0) |  |
| - Men |  |  |  |  |
| 0 units | 36 (60.0)* | 8 (44.4)* | 5 (11.9) | **<0.001** |
| 1-7 units | 23 (38.3) | 9 (50.0) | 27 (64.3) |  |
| 8-14 units | 1 (1.7) | 1 (5.6) | 10 (23.8) |  |
| ≥14 units | 0 (0.0) | 0 (0.0) | 0 (0.0) |  |
| - Women |  |  |  |  |
| 0 units | 10 (55.6)* | 6 (75.0)* | 3 (18.8) | **0.017** |
| 1-7 units | 7 (38.9) | 2 (25.0) | 13 (81.3) |  |
| 8-14 units | 1 (5.6) | 0 (0.0) | 0 (0.0) |  |
| ≥14 units | 0 (0.0) | 0 (0.0) | 0 (0.0) |  |
| Recreational drug abuse, n (%) | 15 (18.8)* | 5 (19.2)* | 0 (0.0) | **<0.001** |
| - Men | 15 (24.6)* | 5 (27.8)* | 0 (0.0) | **<0.001** |
| - Women | 0 (0.0) | 0 (0.0) | 0 (0.0) |  |

### Study program

Eligible participants, both active and previous users, and non-users, were invited to participate in a three-hour examination program carried out at the outpatient clinic at the Department of Cardiology at Odense University Hospital, where all examinations were performed using the same equipment and study protocols.

Upon arrival and after providing written and informed consent, participants were asked about their medical history, including chronic diseases, allergies, and use of prescribed medications. They also provided information about their supplement usage, alcohol and tobacco consumption, drug use, dietary habits, physical activity regimen, and socioeconomic status. Subsequently, participants were asked to provide detailed information on their fitness training history, as well as their experience with illicit AAS, including current or previous use, time intervals between intake, types of AAS, dosage, duration of use, age of onset of AAS use, the maximum weekly dose of AAS, and cumulative lifetime AAS usage. Additionally, participants gave information about their illicit use of other performance-enhancing drugs, such as human growth hormone (hGH), selective estrogen receptor modulators, as well as the use of recreational drugs, including cocaine, cannabis, ecstasy, gamma-hydroxybutyrate (GHB), heroin, lysergic acid diethylamide (LSD), and others. All information was collected through questionnaires.

Participants were grouped according to their self-reported use of AAS. Active users were individuals who reported a current use of AAS, or had discontinued the use of AAS within three months prior to the evaluation. Previous users had discontinued their AAS use at least three months before the study commenced. Non-users had never used AAS.

Urine samples were tested for steroid metabolites in order to verify or dispute the reported use of AAS. Urine analyses of steroid metabolites were conducted at the Centre for Preventative Doping Research at the German Sports University Cologne, Germany. Guidelines for identifying exogenous androgens included the detection of synthetic testosterone compounds in urine samples, and a testosterone/epitestosterone ratio (T/E) exceeding 4 (**32**). Blood samples were collected to measure renal function, lipid levels, and concentrations of testosterone, luteinizing hormone (LH), follicle-stimulating hormone (FSH), and estradiol. These samples were analyzed using standard clinical methods at the Department of Biochemistry, Odense University Hospital, Denmark. In addition, serum and plasma was collected for future studies.

Data on the participants included height, weight, waist and hip circumferences, body mass index (BMI, kg/m²) and handgrip strength (Jamar Hand Dynamometer®) as well as body composition using a bioelectrical impedance spectroscopy apparatus (SOZO®).

All participants underwent bilateral B-mode vascular ultrasound examination of the carotid and femoral arteries, focusing on the presence of plaques, using an ultrasound device (Logiq E, GE, 12 MHz linear transducer). Images were digitally stored and subsequently analyzed. The presence of plaque was defined as a focal structure that extends into the arterial lumen by at least 0.5 mm or 50% greater than the surrounding intima-media thickness (IMT), or an intima-media thickness (IMT) of >1.5 mm. All findings were evaluated by an external validator, who was blinded to the AAS use status of the participants.

Subsequently, a transthoracic echocardiography (GE Vivid E95) was performed, using standard two-dimensional and Doppler echocardiographic views. Recordings from all participants were saved as digital files and analyzed using the software EchoPac^TM^ (GE Healthcare) by a proficient operator blinded to the AAS use status. The following measurements were recorded: cardiac dimensions, LV and RV systolic function, LV diastolic function, and left atrial volume. Systolic function was evaluated using Simpson’s biplane method assessing LV ejection fraction (LVEF) and global longitudinal strain (GLS) analyses. Diastolic function was evaluated by peak mitral inflow velocity (E), early septal and lateral LV relaxation velocity E’, and left atrial volume.

Finally, CCTA (Siemens Force CT scanner), coupled with a semiautomatic computer program (Syngo.Via, Siemens Healthcare) was used to estimate CACs using the Agatston method (**33**). Coronary NCP was visually assessed by an experienced cardiologist. NCP were identified as coronary arterial wall lesions that exhibit low attenuation values relative to the surrounding tissues; for further details please refer to (**31**).

### Statistical analysis

Descriptive statistics were used to summarize demographic and clinical characteristics of the study population. Continuous variables were presented as median (interquartile range) or median (range), and categorical variables were presented as number (n) and percentage (%). The normality of data distribution was assessed using Q-Q-plots and the Shapiro-Wilk test. To address non-normality of the data distributions, transformations were applied.

Comparisons between the three groups were performed using one-way analysis of variance (ANOVA) or Kruskal-Wallis test for continuous variables. Tests for trends across ordered groups were performed using regression analyses or the Jonckheere-Terpstra test. Chi-squared tests were used for categorical variables. The homogeneity of variances assumption was tested using Levene’s test. Associations between variables were analyzed using linear regression.

The effect of cumulative lifetime duration of AAS intake was assessed in the entire cohort using regression models. Linear regression was used to estimate regression coefficients and associated 95% confidence intervals (CIs) for echocardiographic characteristics, and logistic regression was used to estimate odds ratios (OR) and 95% CIs for carotid and femoral plaques, CAC scores, and coronary NCP. The CAC variable was dichotomized (CAC = 0 versus CAC > 0). To control for potential confounding, primary analyses were adjusted in multivariable regression models for co-variables age, sex, body fat percentage, hours of exercise and strength training, family history of coronary disease, blood pressure, total-cholesterol, smoking status, alcohol consumption, and use of recreational drugs. Missing categorical variables (hours of exercise and strength training, family history of coronary disease, blood pressure, cholesterol, smoking status, and alcohol consumption) were imputed on a continuous scale and rounded to the nearest integer. The imputed models were validated by comparing the mean (SD) of the imputed data set with the original data set. Missing data did not exceed 5% for any imputed variables.

All statistical tests were two-sided, and a p-value <0.05 was considered statistically significant. Statistical analyses were performed using Stata software version 18 (StataCorp LP, College Station, TX, USA).

### Power calculation

We aimed to describe a broad group of Danish recreational athletes with at least 3 months of AAS use, targeting a highly diverse population between 18 to 65 years, including both men and women, with a duration of AAS ranging from 3 months to several years. This approach was chosen to enhance the generalizability of our findings, identify potential subgroups, and ensure robustness and variability in the results. Given this anticipated heterogeneity, we refrained from performing a strict power calculation.

Instead, we aimed to include an appropriate number of subjects: targeting 120 active or previous AAS users and 60 age- and sex-matched non-users, as reported previously (**31**).

## RESULTS

### Cohort characteristics

We managed to enroll 80 active users (19 women), 26 previous users (8 women), and 58 healthy non-users (16 women) (**Table 1**). One previous AAS using man was excluded due to discrepancies between his self-reported cessation in 2017 and the urine analysis at enrolment, which detected metenolone and dehydrochloro-methyltestosterone (DHCMT) at low but detectable levels, along with a suspiciously high testosterone/epitestosterone ratio.

No differences in age or sex distribution were observed among the three groups (**Table 1**). Active users had significantly higher BMI, lower fat percentages and higher muscle percentages, followed by previous users and non-users. These trends were consistent across both sexes (**Table 1**).

The duration of AAS use ranged from 3.6 months to 42 years in active users and from 3.6 months to 24.5 years in previous users. In both groups, the median cumulative lifetime duration of AAS intake was 2.2 years (**Table 1**). However, male athletes had a longer duration of AAS use than females, irrespective of whether they were active users or previous users (**Table 1**). Distribution of cumulative years of AAS use among male and female AAS users is illustrated in **Supplementary** Figure 1. Previous users reported discontinuing AAS intake 2.5 years (range 3 months to 29 years) before the investigation date, with no difference between men and women. The year of cessation of AAS intake in is shown in **Supplementary** Figure 2. The type of AAS differed between men and women (p<0.001) (**Figure 1**). Testosterone emerged as the overwhelmingly preferred choice among men during the peak dosage period (73%), whereas women primarily used oxandrolone (43%). Creatinine, hGH, and weight-losing agents turned out as the most frequently used substances alongside AAS **(Supplementary** Figure 3).

**Figure 1.**
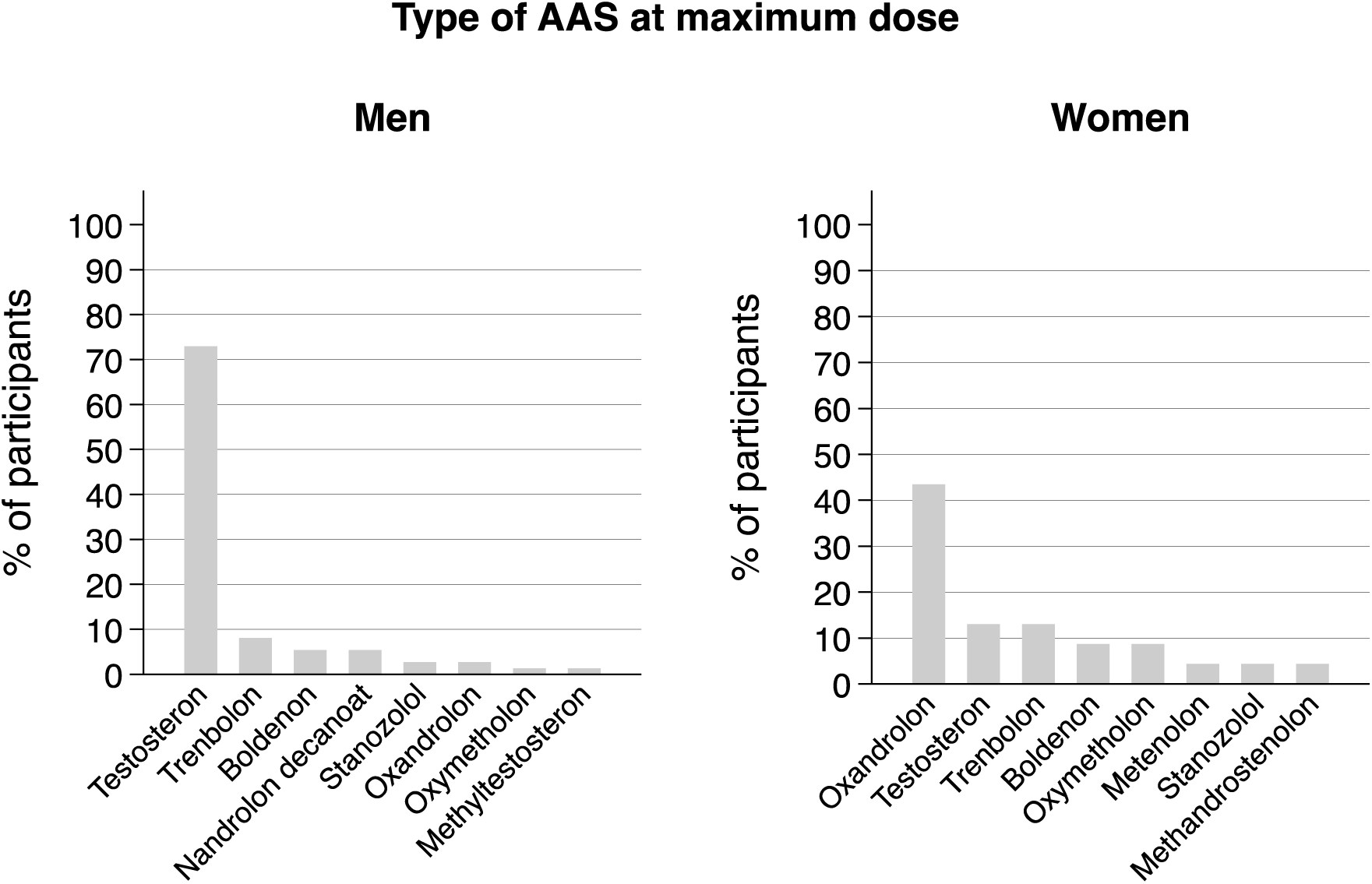
Types of AAS used by active and previous AAS users at the maximum dose in men (left panel) and women (right panel). AAS, androgenic anabolic steroids.

Systolic blood pressure was elevated in active AAS users compared to previous users and non-users, while diastolic blood pressure was lowest in previous users. These findings were reproduced when stratifying according to sex and most likely reflect that previous AAS users received more antihypertensive medication than both active users and non-users **(Supplementary Table 1)**. Finally, both active and previous AAS users showed higher rates of mental disorders such as ADHD and anxiety compared to non-users **(Supplementary Table 1)**.

Regarding lifestyle, active AAS users had higher prevalence of current smokers and fewer never-smokers compared to the non-user group (**Table 1**). However, difference in smoking habits was only evident in men. In contrast, both active and previous AAS users reported significantly lower alcohol consumption than non-users, whereas the use of recreational drugs, primarily cocaine and cannabis was more common among active and previous AAS users than non-users, and apparently limited to men only.

### Biochemistry

Active AAS users showed higher hematocrit and creatinine levels compared to previous users and non-users, with previous users having higher levels than non-users (**Table 2**). In contrast, HbA1c and estimated average glucose levels were similar across the groups. Active AAS users had lower HDL cholesterol and higher LDL cholesterol compared to previous users, while previous users and non-users had similar cholesterol levels. Triglyceride levels did not differ among the groups. These metabolic changes were consistent when analyzing men and women separately.

**Table 2.**
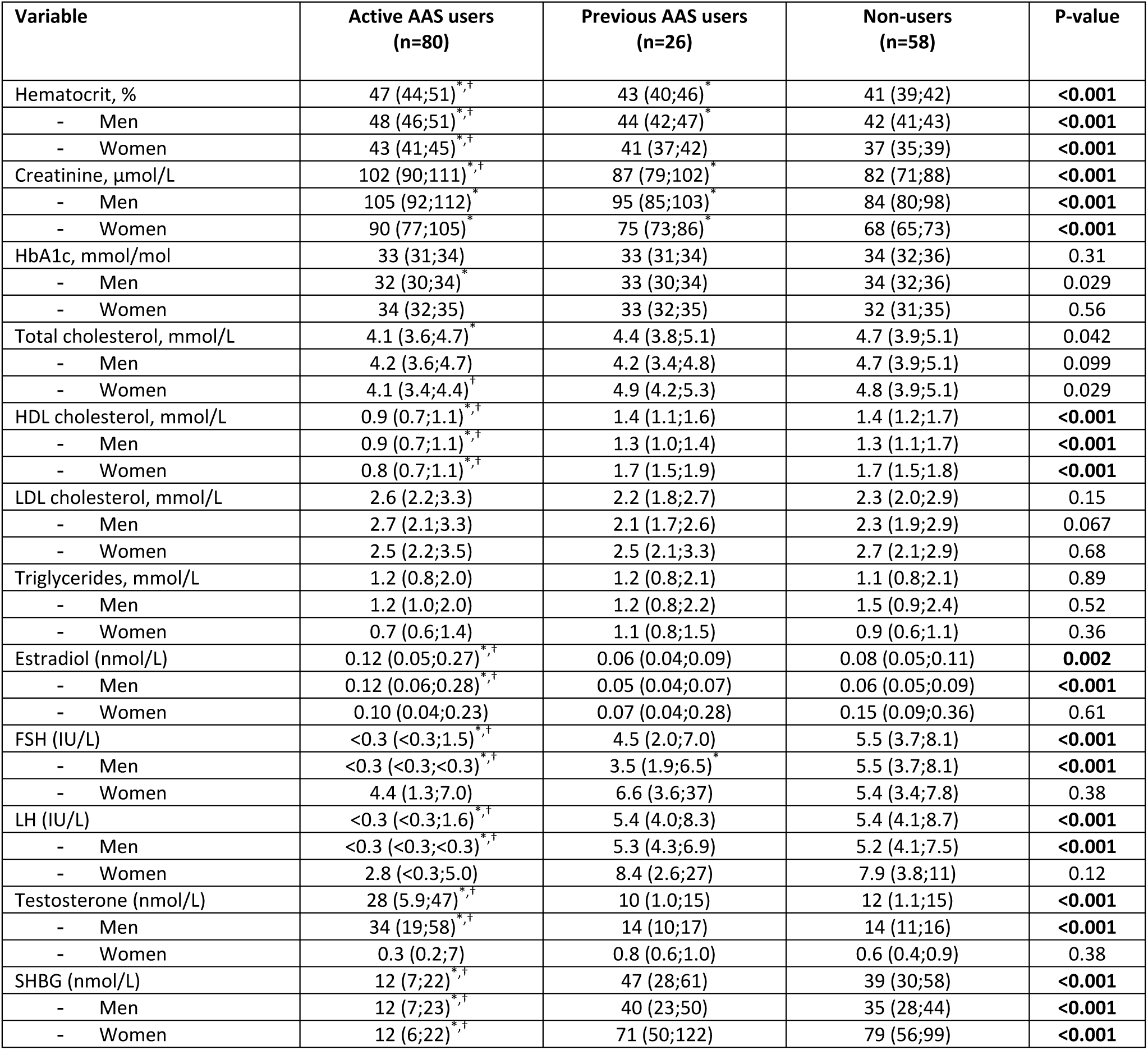
Biochemical characteristics Biochemical characteristics of the participants in the entire cohort stratified by AAS use. Values for continuous variables are shown as median (interquartile range). *p<0.05 when compared to non-users, ^†^p<0.05 when compared to previous AAS users. AAS, androgenic anabolic steroids; FSH, follicle-stimulating hormone; HbA1c, glycated hemoglobin; HDL, high-density lipoprotein; LDL, low-density lipoprotein; LH, luteinizing hormone; SHBG, sex hormone binding globulin.

Regarding sex hormone levels (**Table 2**), active male AAS users demonstrated gonadal suppression, as indicated by low levels of FSH and LH, and increased estradiol levels. Previous male users exhibited significantly lower FSH levels compared to non-users, while LH levels did not show a significant difference.

### Atherosclerosis (carotid, femoral and coronary artery plaques)

One previous AAS using man (age 63 at enrolment) suffered from a non-ST elevation myocardial infarction requiring stenting at the age of 52 years, following 10 years of cumulative AAS intake. None of the non-users had any medical history of CAD.

Active users, previous users, and non-users were compared as regards ultrasound findings (plaques in the femoral or carotid artery) and coronary CT angiography (CAC scores and NCP). With one exception, the statistical analysis did not reveal any significant differences between the three groups (**Table 3**), whether analyzing the entire population or stratifying by sex. The only significance was observed when comparing the fraction of coronary NCP across the entire cohort of men and women, but this did not translate into detectable differences between the groups.

**Table 3.** Carotid and femoral artery plaques by ultrasound, and non-calcified plaques and calcified plaques (CAC score) by CT Results from ultrasound of the femoral artery and the carotid, and coronary CT scans of the participants in the entire cohort stratified by AAS use. CAC scores are shown as median (range). Categorical variables are indicated as number (n) and percentage (%) of persons. *p<0.05 when compared to non-users, ^†^p<0.05 when compared to previous AAS users. AAS, androgenic anabolic steroids; CAC, coronary artery calcium; NCP, coronary non-calcified plaque.

| Dependent variable | Active AAS users<br>(n=80) | Previous AAS<br>users<br>(n=26) | Non-users<br>(n=58) | P-value<sub>groups</sub> | P-value<sub>gender</sub> |
| --- | --- | --- | --- | --- | --- |
| NCP, n (%) | 19 (23.8) | 4 (15.4) | 6 (10.3) | **0.041** |  |
| - Men | 16 (26.2) | 4 (22.2) | 5 (11.9) | 0.082 | 0.11 |
| - Women | 3 (15.8) | 0 (0.0) | 1 (6.3) | 0.321 |  |
| CAC score | 0 (0;228) | 0 (0;800) | 0 (0;163) | 0.355 |  |
| - Men | 0 (0;228) | 0 (0;800) | 0 (0;163) | 0.759 | 0.26 |
| - Women | 0 (0;166) | 0 (0;0) | 0 (0;2) | 0.166 |  |
| Femoral artery plaque, n (%) | 15 (19.7) | 5 (19.2) | 11 (20.8) | 0.894 |  |
| - Men | 12 (20.7) | 4 (22.2) | 9 (23.7) | 0.729 | 0.32 |
| - Women | 3 (16.7) | 1 (12.5) | 2 (13.3) | 0.784 |  |
| Carotid artery plaque, n (%) | 49 (31.2) | 12 (46.2) | 13 (24.1) | 0.467 |  |
| - Men | 16 (27.1) | 9 (50.0) | 11 (28.2) | 0.789 | 0.97 |
| - Women | 8 (44.4) | 3 (37.5) | 2 (13.3) | 0.061 |  |

We subsequently evaluated the cumulative lifetime duration of AAS use as an independent predictor for atherosclerosis in the entire population, as well as separately in men and women. Both univariate and multivariable logistic regression analyses revealed that cumulative AAS use was associated with a positive CAC score and the presence of coronary NCP (**Table 4**). The association with carotid artery plaques was of borderline significance (p<0.089) and non-significant for femoral artery plaques. In sex-specific analyses, significant associations were found only in men, specifically with regards to CAC score and coronary NCP (**Table 4**).

**Table 4.** Atherosclerotic stigmata and their relationship with cumulative lifetime duration of AAS use Univariable and multivariable logistic regression analyses of the entire population with cumulative lifetime duration of AAS use as independent predictor for atherosclerotic measures. The CAC variable was dichotomized (CAC = 0 versus CAC > 0). The multivariable model included co-variables age, body fat percentage, family history of coronary artery disease, blood pressure, total-cholesterol, use of recreational drugs, tobacco use, alcohol consumption, and hours of hard exercise and strength training. AAS, androgenic anabolic steroids; CAC, coronary artery calcium; CI, confidence interval; NCP, coronary non-calcified plaque; OR, odds ratio.

| Dependent variable | Cumulative lifetime duration of AAS use, years |  |  |  |
| --- | --- | --- | --- | --- |
|  | Univariable logistic regression<br>OR (95% CI) | P-value | Multivariable logistic<br>regression<br>OR (95% CI) | P-value |
| NCP | 1.17 (1.09;1.26) | <0.001 | 1.17 (1.05;1.30) | **0.004** |
| - Men | 1.18 (1.09;1.28) | <0.001 | 1.21 (1.06;1.38) | **0.005** |
| - Women | 0.78 (0.37;1.65) | 0.512 | 0.53 (0.13;2.26) | 0.39 |
| CAC score | 1.18 (1.09;1.27) | <0.001 | 1.23 (1.09;1.39) | **0.001** |
| - Men | 1.20 (1.10;1.31) | <0.001 | 1.43 (1.17;1.73) | **<0.001** |
| - Women | 0.80 (0.422;1.50) | 0.481 | 0.43 (0.08;2.31) | 0.32 |
| Femoral artery plaque | 1.05 (1.00;1.10) | 0.044 | 0.98 (0.92;1.05) | 0.63 |
| - Men | 1.05 (1.00;1.11) | 0.045 | 0.99 (0.92;1.08) | 0.87 |
| - Women | 0.78 (0.44;1.41) | 0.416 | 0.17 (0.02;1.64) | 0.12 |
| Carotid artery plaque | 1.10 (1.03;1.17) | 0.002 | 1.08 (0.99;1.18) | 0.089 |
| - Men | 1.09 (1.02;1.15) | 0.006 | 1.06 (0.97;1.16) | 0.19 |
| - Women | 1.44 (1.03;2.01) | 0.031 | 1.35 (0.88;2.08) | 0.17 |

Given the association between cumulative AAS use and a positive CAC score, we further stratified the entire AAS user group into subgroups based on increasing durations of use (>1, >2, >3, >4, >5, and >10 years) to identify when the risk of a positive CAC score began to rise (**Figure 2A, left panel)**. After adjusting for age and sex, the risk of a positive CAC score increased after >3 years of AAS use (OR: 3.3, 95% CI: 1.05;10.5), was borderline significant after >4 years (OR: 3.1, 95% CI: 0.96;9.8), and became significant again after >5 (OR: 4.5, 95% CI: 1.22;16.25) and >10 years (OR: 12.0, 95% CI: 2.5;58.3). A similar trend was observed in the fully adjusted model (**Figure 2A, right panel)**, although the OR did not reach statistical significance.

**Figure 2.**
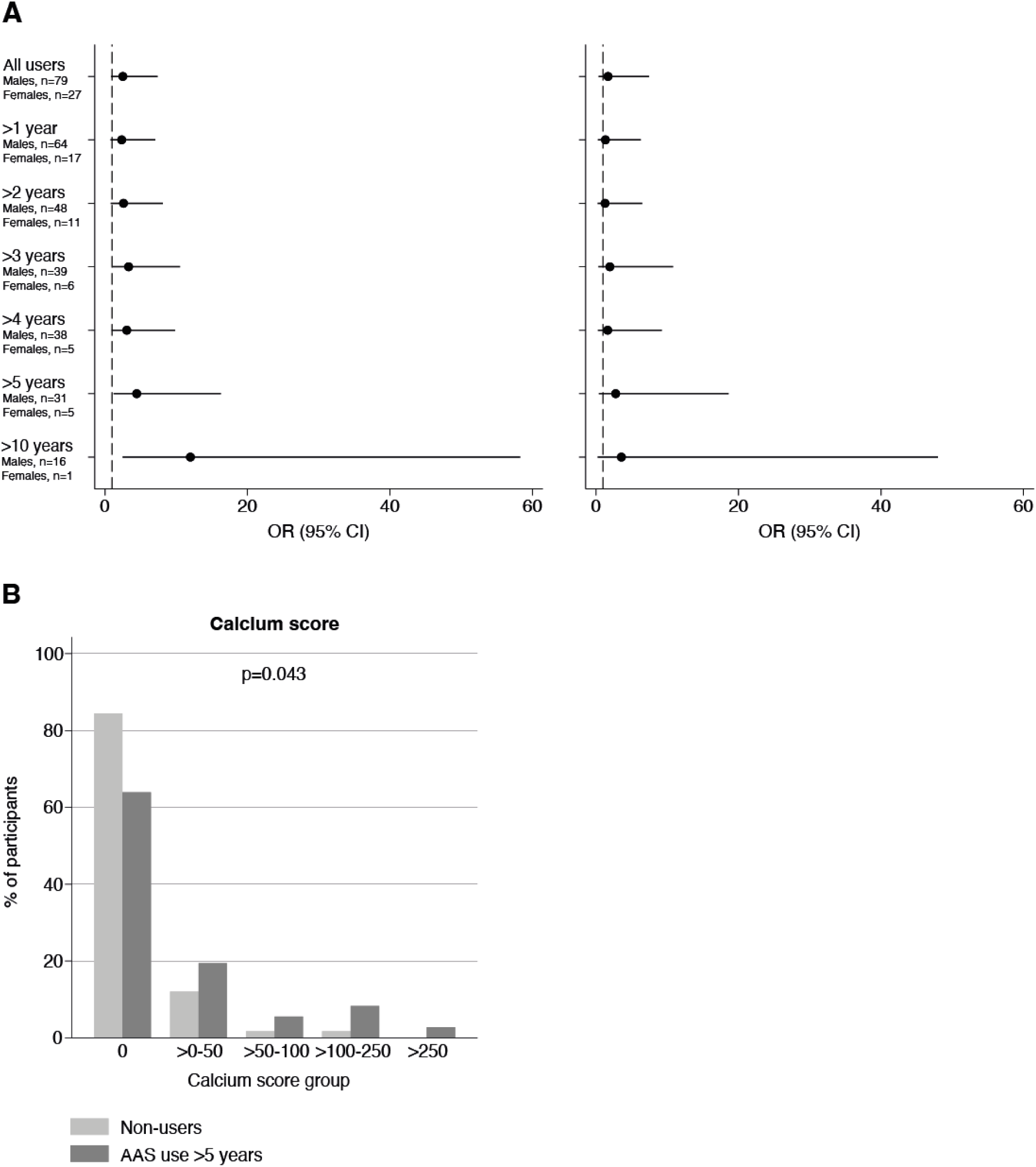
A) Logistic regression analysis comparing the presence (CAC scores > 0) versus the absence (CAC score = 0) of coronary artery atherosclerosis in the 58 non-users versus AAS users (previous and active users) stratified by cumulative lifetime duration of AAS intake. The number of AAS users in each group is shown. Models were adjusted for age and gender (left panel), and age, gender, body fat percentage, family history of coronary artery disease, blood pressure, total-cholesterol, use of recreational drugs, tobacco use, alcohol consumption, and hours of hard exercise and strength training (right panel). B) CAC score in the AAS users with a cumulative AAS intake of >5 years (n=36) vs. non-users (n=58). AAS, androgenic anabolic steroids; CAC, coronary artery calcium; CI, confidence interval; OR, odds ratio.

The logistic regression analysis focused on the presence of a positive CAC score (yes or no) but did not account for the severity of calcification. To address this, we compared the fraction of non-users (n=58) and subjects with a cumulative AAS use >5 years (n=36) when divided into CAC score categories (0; >0 to 50; >50 to 100; >100 to 250; >250), representing increasing atherosclerotic severity. The two groups of non-users and long-term users (> 5 years) did not differ significantly as regards mean age (p=0.47) or sex distribution (p=0.12). The threshold of >5 years of AAS use was based on the logistic regression which suggested that after this period of use, the odds of having a positive CAC score approached significance, and indeed, AAS users with a cumulative use of AAS exceeding 5 years had significantly more severe CAC scores compared to non-users (**Figure 2B**).

### Cardiac function by echocardiography

At the age of 29 years, one active male AAS user was diagnosed with non-ischemic congestive heart failure having a LVEF of 20%. This individual received ACE inhibitor and beta-blocker medication, which improved LVEF to 40%. Nobody else had any history of structural heart disease.

Both active and previous users demonstrated a significantly higher prevalence of altered left ventricular geometry compared to non-users (**Table 5**). In the entire cohort, echocardiography demonstrated numerous significant differences in both systolic and diastolic function, with changes in active and previous users compared to non-users (**Table 5**). LVEF displayed a gradual decline, with active AAS users showing the lowest values compared to previous AAS users and non-users. The same pattern was evident for LV GLS, and RV GLS. Moreover, when evaluating ventricular relaxation, active AAS users exhibited higher E/E’ ratios compared to previous users and non-users. These echocardiographic findings were consistent across both sexes **(Supplementary** Figure 4), with men and women displaying similar changes in LVEF, LV and RV strains, and E/E’ ratios among active users, previous users, and non-users. In contrast, left atrial volume did not differ in either men or women (**Table 5**).

**Table 5.** Echocardiographic findings Echocardiographic characteristics of the participants in the entire cohort stratified by AAS use. Values for continuous variables are shown as median (interquartile range). Categorical variables are indicated as number (n) and percentage (%) of persons. *p<0.05 when compared to non-users, ^†^p<0.05 when compared to previous AAS users. AAS, androgenic anabolic steroids.

| Variable | Active AAS users (n=80) | Previous AAS users (n=26) | Non-users (n=58) | P-value |
| --- | --- | --- | --- | --- |
| Left ventricular ejection fraction, % | 54 (52;56)<sup>*,†</sup> | 57 (55;58) | 59 (56;61) | <0.001 |
| - Males | 54 (51;56)<sup>*,†</sup> | 57 (54;58) | 59 (56;60) | <0.001 |
| - Females | 54 (53;56)<sup>*,†</sup> | 57 (55;59) | 59 (57;62) | <0.001 |
| Left ventricular global longitudinal strain | -18 (-19;-17)<sup>*,†</sup> | -19 (-20;-18) | -20 (-21;-19) | <0.001 |
| - Males | -18 (-19;-17)<sup>*,†</sup> | -19 (-20;-18) | -20 (-20;-18) | <0.001 |
| - Females | -18 (-19;-17)<sup>*,†</sup> | -19 (-19;-18) | -20 (-22;-19) | <0.001 |
| Right ventricular global longitudinal strain | -20 (21;-20)<sup>*,†</sup> | -21 (-22;-21) | -23 (-24;-21) | <0.001 |
| - Males | -20 (-21;-20)<sup>*,†</sup> | -21 (-23;-21) | -22 (-24;-21) | <0.001 |
| - Females | -20 (-21;-20) | -21 (-22;-21) | -23 (-25;-22) | <0.001 |
| Early mitral peak velocity, cm/s | 72 (68;77) | 71 (64;78) | 74 (68;85) | 0.13 |
| - Males | 71 (67;76) | 69 (63;74) | 72 (64;81) | 0.67 |
| - Females | 74 (68;87) | 72 (68;79) | 85 (76;94) | 0.053 |
| Early left ventricular relaxation (septal) E', cm/s | 9 (9;10) | 10 (9;11) | 12 (11;13) | <0.001 |
| - Males | 9 (9;10) | 10 (9;11) | 12 (10;13) | <0.001 |
| - Females | 10 (9;11) | 11 (10;12) | 13 (12;13) | <0.001 |
| Early left ventricular relaxation (lateral) E', cm/s | 12 (11;13) | 13 (12;14) | 15 (14;16) | <0.001 |
| - Males | 12 (11;13) | 12 (11;14) | 15 (14;16) | <0.001 |
| - Females | 13 (12;14) | 14 (13;15) | 16 (15;17) | <0.001 |
| Early mitral peak velocity and average left ventricular relaxation (E/E') ratio | 7.0 (6.3;7.8)<sup>*,†</sup> | 6.4 (5.7;6.8) | 5.8 (5.1;6.3) | <0.001 |
| - Males | 7.1 (6.3;7.8) | 6.6 (6.0;6.9) | 5.6 (5.0;6.2) | <0.001 |
| - Females | 6.7 (6.2;7.4)<sup>*,†</sup> | 5.8 (5.7;6.4) | 6.3 (5.6;6.5) | 0.013 |
| Left ventricular mass, g | 194 (164;230) | 176 (153;234) | 144 (123;170) | <0.001 |
| - Males | 207 (182;242) | 194 (164;234) | 158 (142;178) | <0.001 |
| - Females | 159 (148;176)<sup>*,†</sup> | 135 (119;170) | 106 (96;123) | <0.001 |
| Left ventricular mass/body surface area, g/m2 | 93 (85;105) | 85 (77;111) | 68 (62;77) | <0.001 |
| - Males | 97 (86;107) | 97 (79;117) | 71 (67;82) | <0.001 |
| - Females | 86 (83;99)<sup>*,†</sup> | 82 (64;91) | 58 (51;65) | <0.001 |
| Relative wall thickness | 0.42 (0.38;0.45) | 0.41 (0.37;0.43) | 0.36 (0.32;0.39) | <0.001 |
| - Males | 0.42 (0.39;0.45) | 0.42 (0.40;0.46) | 0.37 (0.33;0.39) | <0.001 |
| - Females | 0.41 (0.38;0.45)<sup>*,†</sup> | 0.34 (0.31;0.38) | 0.32 (0.31;0.37) | <0.001 |
| Left atrial volume, ml | 43 (40;47) | 42 (38;47) | 44 (41;49) | 0.19 |
| - Males | 45 (42;48) | 44 (39;48) | 47 (42;51) | 0.084 |
| - Females | 38 (38;40) | 38 (36;40) | 39 (35;40) | 0.72 |
| Cardiac remodeling and hypertrophy, n (%) |  |  |  |  |
| Normal geometry | 40 (50.0)<sup>*</sup> | 16 (61.5)<sup>*</sup> | 51 (87.9) | <0.001 |
| Remodeling | 28 (35.0) | 6 (23.1) | 7 (12.1) |  |
| Hypertrophy | 12 (15.0) | 4 (15.4) | 0 (0.0) |  |
| - Males |  |  |  |  |
| Normal geometry | 28 (45.9)* | 9 (50.0)* | 36 (85.7) | **<0.001** |
| Remodeling | 24 (39.3) | 5 (27.8) | 6 (14.3) |  |
| Hypertrophy | 9 (14.8) | 4 (22.2) | 0 (0.0) |  |
| - Females |  |  |  |  |
| Normal geometry | 12 (63.2) | 7 (87.5) | 15 (93.8) | 0.23 |
| Remodeling | 4 (21.1) | 1 (12.5) | 1 (6.3) |  |
| Hypertrophy | 3 (15.8) | 0 (0.0) | 0 (0.0) |  |

Multivariate analysis revealed several significant associations between cumulative lifetime AAS use and various echocardiographic measures **(Supplementary Table 2)**. Participants with prolonged AAS use exhibited more pronounced cardiac impairments. **Figure 3** highlights the relationship between cumulative AAS intake and key echocardiographic parameters—LVEF, LV mass, E/E’ average, and GLS—showing that nearly all athletes with more than 5 years of AAS use had values outside the median of the normal range for these measures.

**Figure 3.**
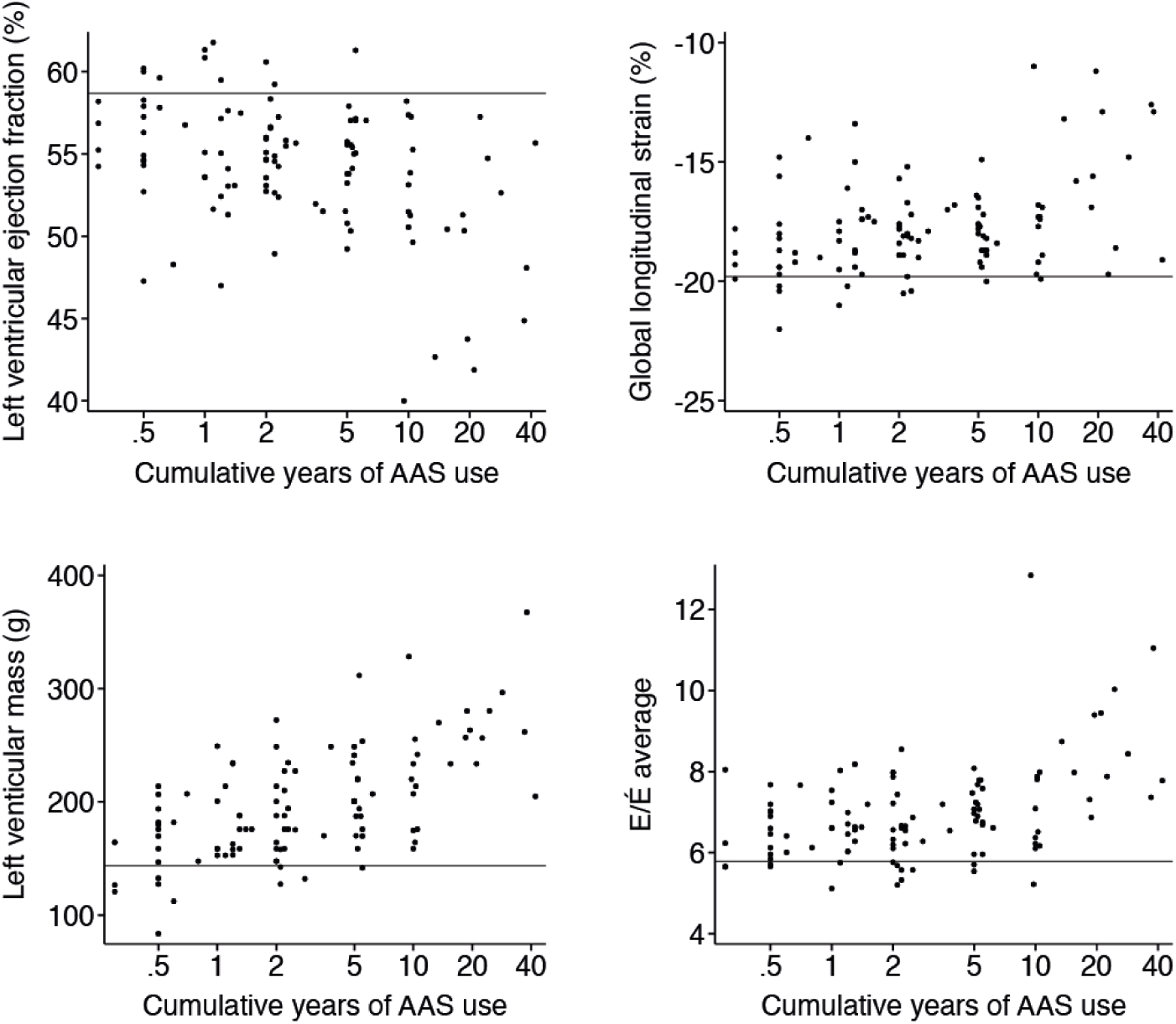
Relationships between echocardiographic parameters and the cumulative years of AAS intake in active and previous AAS users. The horizontal lines represent medium levels in the non-user group. AAS, androgenic anabolic steroids.

## DISCUSSION

Our understanding of the cardiovascular consequences of illicit AAS use is primarily derived from case reports (**7, 8, 10, 12, 13**), minor observational studies (**9, 29**), and epidemiological data (**6**), predominantly involving men in specific athletic cohorts. To address this gap, this study was designed to provide new insights into a broader population of AAS-using recreational athletes, including women, for whom data are virtually nonexistent. Data was successfully collected from 106 recreational athletes, including 27 women. Although the male-to-female ratio in our study may be coincidental, it roughly aligns with estimates that AAS use is approximately four times less common among women compared to men (**3**).

Using vascular ultrasound and CT coronary angiography, preclinical markers of cardiovascular disease was assessed by comparing active users, previous users, and non-users. No significant differences in the prevalence of femoral artery plaques, carotid artery plaques, CAC scores, or coronary NCP between AAS users and non-users was found (**Table 3**). This lack of difference may be due to the inclusion of many younger athletes with short-term AAS use. However, univariate and multivariable logistic regression analyses identified cumulative lifetime AAS use as a significant and independent predictor of both calcified and non-calcified coronary artery plaques, particularly in the whole cohort and in men. This highlights that the lifetime duration of AAS exposure, rather than current user status (previous or ongoing), is critical in assessing the risk of premature atherosclerotic disease. Of notice, these data align with observations by Baggish *et al.* (**15**), who linked lifetime AAS exposure to coronary atherosclerosis in experienced male weightlifters. In women, neither soft nor hard coronary artery plaques were associated with lifetime AAS use, likely due to the smaller sample size and shorter duration of AAS use compared to men. However, it cannot be ruled out that this may also reflect the female biological advantages in relation to cardiovascular disease development (**34**).

When stratifying the cohort by lifetime AAS exposure, logistic regression analysis revealed that, after adjusting for age and sex, an AAS lifetime duration of more than 3 years was significantly associated with an increased likelihood of a positive CAC score. This association was of borderline significance at 4 years but regained significance for AAS users with a lifetime duration of more than 5 and 10 years. However, when the model was fully adjusted for additional risk factors, the associations were no longer significant, though the trends remained consistent with the lesser-adjusted model (**Figure 2, upper panels)**. This suggests that traditional cardiovascular risk factors, such as lifestyle, also play an important role in illicit AAS users.

Since the logistic regression analysis focused on comparing the presence (CAC scores > 0) versus the absence (CAC score = 0) of coronary artery atherosclerosis without accounting for the severity of calcifications, we further investigated the relationship between the severity of coronary artery calcifications and the lifetime duration of AAS use. Using a cut-off of more than 5 years of AAS intake, a fully adjusted model (**Figure 2, lower panel)** was applied and it demonstrated that a cumulative AAS use exceeding 5 years is associated with an increased CAC score. Again our data align well with those reported by Baggish *et al.* (**15**) and suggest that findings from studies of selected AAS-using athletes may also be applicable to a broader population of recreational athletes.

We assessed cardiac function and found that both men and women exhibited similar declines in LV and RV systolic function, as well as LV diastolic function. Analysis stratified by sex revealed consistent adverse effects of AAS on cardiac function across genders. A decline in RV systolic function was observed, indicating a global impact on systolic performance. This finding extends beyond previous research, which predominantly focused on LV systolic function (**16, 26, 29**). Notably, echocardiographic findings indicated that these changes can occur early in the course of AAS use and do not require extended exposure. Another finding was that after 5 years of intake of AAS, virtually all AAS users had echocardiographic findings that were outside normal range (**Figure 3**). In conjunction with the CAC score data this indicates that 5 years of exposure to AAS constitutes a threshold, where-after the odds of having increased coronary calcification and a subnormal echocardiography is significantly elevated as compared to non-users. On the other hand, the echocardiographic comparison between previous and active AAS users indicated that myocardial dysfunction was less prevalent in those who had ceased AAS use, suggesting that discontinuation may reduce some of the risks of lasting cardiac damages.

Overall, the trends observed when comparing characteristics of athletes with active AAS use, previous AAS use, and non-users were similar between men and women, though fewer significant differences were found among the female groups, likely due to smaller sample sizes (**Table 1**). In terms of physical training, women spent as many hours in the gym as men, regardless of their AAS use status. Additionally, both sexes spent twice as many hours per week in the gym as non-users. However, despite this, neither hand-grip strength nor self-reported maximum bench-press weight was higher in active AAS-using women compared to non-users, while men with ongoing AAS use were significantly stronger than previous users and non-users. It remains unclear whether these differences are due to variations in training programs or the use of AAS and other performance-enhancing drugs.

Our study also revealed notable differences in lifestyle factors between women and men (**Table 1**). Both active and previous AAS users of both sexes reported lower alcohol intake compared to non-users.

However, men actively using AAS reported a lower proportion of never smokers, with a similar trend observed among women. The reduced alcohol intake among active and previous AAS users may indicate a prioritization where alcohol, but not cigarettes, is viewed as detrimental to physical performance.

Conversely, the use of recreational drugs (primarily cocaine and cannabis), both established cardiovascular risk factors (**35–37**), was reported exclusively by men. The sex difference suggests that men with active or previous AAS use are more prone to risk-taking behaviors than women, indicating a potential difference in risk behavior between male and female AAS users.

AAS intake led to similar biochemical changes in both men and women (**Table 2**). In both active AAS-using groups, hematocrit levels were elevated, HbA1c remained within the normal range, and HDL levels were lower compared to sex-matched non-users, indicating that AAS intake adversely affects cardiovascular risk markers in both men and women. Endocrine changes in men were as expected (**38**), with FSH and LH being suppressed in men only. Interestingly, women on AAS had normal FSH, LH, testosterone and estrogen levels but significantly suppressed SHBG levels. It is unclear whether this suppression was due to a voluntary pause in AAS intake before the study or the use of testosterone preparations undetected by our assay. However, urinary measurements confirmed that both women and men with active AAS use were accurately categorized.

### Limitations

This study has limitations to consider. First, its cross-sectional design prevents us from establishing causality between illicit AAS use and adverse cardiovascular outcomes. Second, although our enrollment criteria were broad, there is a risk of selection bias. Thus, it cannot be concluded that our cohort fully represents the broader population of Danish recreational athletes with an illicit use of AAS. However, the male-to-female ratio of AAS users in our study aligns with existing estimates (**3**), which is reassuring. Third, self-reported data on AAS use, including types, doses, cycles, and routes of administration, may be unreliable due to recall bias over many years. Therefore, we found it prudent to validate AAS use status (current/previous) through urinary steroid assays, which confirmed user status and highlighted the importance of such validation in doping studies. Finally, we initially intended to measure coronary NCP volumes in coronary arteries but faced challenges due to variations in muscle mass affecting image quality. As a result, we focused on reporting non-calcified plaques (present/absent) and CAC scores, reflecting the possible difficulties in imaging highly muscular AAS users.

## CONCLUSION

Cumulative lifetime AAS intake in recreational athletes independently predicted both soft and hard coronary plaques. We could not statistically identify a specific time point where the cumulative intake of AAS turned out to be an independent predictor for an increased risk of coronary artery calcifications, but our data suggest that a cumulative use of AAS exceeding 5 years constitutes a threshold.

Coronary changes were more common in men. Most likely, this reflects the smaller number of female athletes, sex differences in the use of AAS preparation and doses, or the shorter duration of AAS intake among women. However, we cannot exclude the presence of a genuine sex difference as regards the susceptibility to the harmful effect of AAS. On the other hand, significant associations were found between AAS use and myocardial changes on echocardiography in both sexes. Cardiovascular risk markers, including increased hematocrit, altered lipid profiles, and elevated blood pressure, were also present in both men and women with a history of AAS use. Finally, risk-taking behavior was more prevalent among male AAS users. In summary, our study show that preventive measures against the use of AAS in recreational sports should target both women and men.

## Data Availability

The datasets generated and analyzed during the current study are not publicly available due to privacy and ethical considerations but are available from the corresponding author upon reasonable request. Access will be provided in compliance with applicable regulations and after approval from the relevant institutional review board.

